# Age-specific immunity to rotavirus infection and the risk of disease before and after rotavirus vaccine introduction in the United Kingdom: an observational, seroepidemiological study

**DOI:** 10.1101/2025.04.03.25324959

**Authors:** Daniel Hungerford, Virgina E. Pitzer, Khuzwayo C. Jere, Marc Y R Henrion, Jonathan Mandolo, Catherine Beavis, Karen Ryan, Jenna Lowe, Nigel A. Cunliffe, Neil French, Miren Iturriza-Gómara

## Abstract

**Background:** Infant rotavirus vaccination was introduced in the UK in July 2013, and was followed by a rapid reduction in rotavirus disease. We compare immunity to rotavirus in sera of age-stratified cohorts to assess the effect of vaccination on natural exposure to rotavirus.

**Methods:** Residual serum samples were selected from a reference laboratory biobank based on age, and year of collection. Anti-rotavirus Immunoglobulin G (IgG) and IgA antibodies were measured using ELISA. We used censored linear spline regression models and anti-rotavirus IgA correlate of protection (CoP) thresholds to estimate immunity by age and vaccine eligibility (defined by date of birth).

**Results:** We analysed serum from 1,200 individuals obtained between 2000 and 2019, of which 807 were vaccine-ineligible and 393 were vaccine-eligible. Among infants aged 3-11 months, a seven-fold increase in IgG levels was observed in the vaccine-eligible cohort (Geometric Mean Concentration [GMC], 1388.67 U/mL; 95% confidence interval [CI], 675.03-2856.78) compared to the vaccine-ineligible cohort (GMC, 198.54 U/ml; 95% CI, 71.31-552.78). Models showed increasing IgG and IgA antibody concentrations in the vaccine-ineligible cohort through childhood and into adulthood, but not in the vaccine-eligible cohort. The proportion of children <7 years with IgA≥160 U/ml, a proxy CoP against rotavirus disease, was lower among the vaccine-eligible population (adjusted odds ratio, 0.66; 95% CI, 0.48–0.91).

**Conclusions:** The introduction of rotavirus vaccination has reduced rotavirus disease burden but has disrupted the subsequent acquisition of naturally-acquired immunity. The impact of this on susceptibility to rotavirus disease in later life remains to be determined.

## Background

In July 2013 the UK introduced rotavirus vaccination into it’s routine infant immunisation schedule. The Rotarix® (GlaxoSmithKline Biologicals, Belgium) two-dose monovalent live-attenuated oral rotavirus vaccine is given at 8 and 12 weeks of age. Vaccine uptake in England rapidly reached >90% for one dose.[1] Prior to vaccine introduction, rotavirus was estimated to be responsible for more than 750,000 diarrhoea episodes and 80,000 GP consultations in the UK each year, accounting for 45% of acute gastroenteritis (AGE) hospitalisations in children under 5 years old.[2,3]

National vaccination rapidly resulted in a greater than 80% reduction of rotavirus hospitalisations, with substantial reductions in healthcare use for AGE across primary and secondary care.[4–6] However, the long-term impact of vaccination is not fully understood, with a study from the USA suggesting that there may be a bounce-back of disease in older vaccinated children.[7] Furthermore, modelling suggests that whilst there will continue to be overall reductions in severe rotavirus AGE incidence over time, incidence may actually increase in school-age children and adults in the longer term, depending on the level of vaccine-induced immunity.[8] We therefore need to understand how protection provided by vaccination compares to that of natural rotavirus exposure and how this impacts the risk of rotavirus disease from birth to adulthood.

The incorporation of serological data as an indicator of exposure to rotavirus re-infections over time will help predict how vaccine-induced changes in immunity in the vaccinated population affect disease incidence in school-age children and adults. Immunoglobulin G (IgG) is the most reliable and consistent marker for seroconversion after rotavirus infection and is associated with protection against severe disease.[9–11] Concentrations of IgG specific to rotavirus antigen can therefore be used to estimate the level of exposure to rotavirus infection. Immunoglobulin A (IgA) is an antibody that plays a crucial role in the immune function of mucous membranes, including the gastrointestinal tract. Levels of anti-rotavirus serum IgA are currently used as a marker for seroconversion after vaccination and as a proxy correlate of protection (CoP) from rotavirus disease, with a value of 20 U/ml indicated as a threshold for protection against severe rotavirus disease in high income countries (HICs).[12–14] There has, however, been considerable debate as to whether there are better CoPs than anti-rotavirus serum IgA.[15]

We used anti-rotavirus serum immunoglobulin responses to estimate changes in exposure to rotavirus infection, as a population ages in a vaccine-ineligible and vaccine-eligible population in the UK.

## Methods

### Study design and participants

Serum samples collected between 2000-2019 were obtained from the UK Health Security Agency (UKHSA) Seroepidemiology Programme based at the Seroepidemiology Unit (SEU), Manchester.[16,17] The samples comprise residual sera from routine diagnostic testing, supplied to SEU by diagnostic laboratories in England.

The samples exclude immunocompromised individuals, but the clinical indication for the original blood sample collection is unknown. The samples are anonymised prior to SEU archiving, while retaining data on age, sex, date of collection and source laboratory, and therefore individual vaccination status is unknown.[17]

A total of 1200 serum samples were requested, stratified by age group and sample year. Sample sizes were pragmatic based on available resources but also informed by the below criteria. The number of samples from infants up to 2 years of age was informed by IgA responses to rotavirus vaccination (Rotarix) in Europe at 1-2 months after second dose and age-specific IgG and IgA levels from a seminal pre-vaccine seroepidemiological study in Brazil.[11,18,19] For those aged 0-2 months, we expect no difference in GMC in the pre-vaccine and post-vaccine period. In those aged 3-11 months, we expect greater than 4-fold differences in GMC; therefore, the sample size required is smaller. From the age of 12-23 months and upwards, sample size was based upon detecting an at least 2.5-fold change in IgA and IgG GMC, assuming 80% power and 5% alpha for two-tailed comparison between samples from the vaccine period and pre-vaccine period. Without robust data from HICs, from 2 years of age, the sample size for each age stratum was based upon data from Brazil.[11]

### Ethical approvals

Ethical approval for the seroepidemiology collection at SEU was granted by the NHS Joint UCL/UCLH Committees on the Ethics of Human Research Ethics Committee (REC) (REC reference number, 05/Q0505/45) for the use of the samples for surveillance and research that informs the National Immunisation Programme for England. This study was approved by the UKHSA SEU Steering Committee.

### Patient and public involvement

Patients and the public were consulted through our institute Patient and Public Involvement and Engagement panel and community workshops with parents/caregivers. Parents/caregivers had concerns about whether vaccines provided good protection and wanted access to basic statistics on vaccine effects. Members were involved in the design of this study and development of the grant application and a member of the panel (CB) is a co-author. Study findings were shared in lay form through an NIHR conference with attendance from public contributors. Feedback on the limitations and implications for families are incorporated within this manuscript.

### Laboratory methods

Anti-rotavirus IgG and IgA concentrations were measured using standardised sandwich enzyme-linked immunosorbent assay (ELISA) as previously described.[20] Serological analysis was conducted at the University of Liverpool. Serum samples were transferred from the SEU on dry ice and then stored at −80°C. Briefly, 96-well plates (Costar) were coated with rabbit hyperimmune serum to rotavirus (at a dilution of 1:10,000 for rotavirus-IgA and 1:2000 for rotavirus-IgG), were incubated with purified cell culture lysates (WC3, dilution of 1:2 for rotavirus-IgA and 1:1 for rotavirus-IgG) or mock-infected MA104 cells (dilution of 1:2 for rotavirus-IgA and 1:1 for rotavirus-IgG). Two-fold serial dilutions, starting at 1:20 for IgA and 1:100 for IgG, of standard and test sera were added followed by biotinylated rabbit anti-human IgA (Jackson ImmunoResearch Laboratories, dilution of 1:3000) for detection of rotavirus-IgA and biotinylated rabbit anti-human IgG (Vector Laboratories, dilution of 1:800). Absorbance was subsequently read at 492 nm. Background-corrected optical density values from sample wells were compared with the standard curve, and concentration was determined based on derived units of IgA or IgG arbitrarily assigned to the respective standard curve. For this assay, seroconversion was defined as detection of rotavirus-IgA at ≥201IU/ml in previously seronegative infants or a 4-fold increase in rotavirus-IgA concentration among infants who were seropositive at baseline. The lower limit of quantification (LLoQ) on standard curve for the IgG and IgA assays were 97.1 U/ml and 4.5 U/mL, respectively.[20] Any results above the upper limit of quantification of the standard curve were repeated with further serial dilutions.

### Statistical analysis

Samples were assigned vaccine eligibility status based on birth month. Those born before May 2013 were considered vaccine-ineligible and those born from May 2013 were vaccine-eligible. Anti-rotavirus IgG and IgA concentrations were described by *a-priori* selected age groups and vaccine eligibility, and age-related serological profile charts were generated with geometric mean concentrations (GMC) within each age group.

To formally assess the relationship between antibody levels and age, we used censored piecewise linear regression models for logarithm-transformed IgG/IgA GMC. Censored regression techniques were used because the assays have LLoQ and therefore the data are left-censored with several values below the LLoQ. We also used a linear spline for age, with statistical inference used to select the changepoints (knot locations). This methodology was used by Swarthout et al., 2022,[21] to assess antibody levels induced by a 13-valent pneumococcal conjugate vaccine and is described on GitHub.[22] We fitted separate models for rotavirus-specific IgA and IgG by vaccine eligibility status; starting values were supplied for up to three knot locations and the optimal number of knots selected using the Akaike information criterion (AIC). Bootstrap resampling and percentile methods were used to calculate 95% confidence intervals for the slope, knot location, age-group-specific GMC, and model-estimated concentrations. Model and GMC plots are log-linear scale with log y-axis, so model fit is presented linearly.

As a post hoc analysis, we also calculated the proportion of samples tested for anti-rotavirus IgA with ≥20 U/ml as a CoP against severe disease and two thresholds of ≥160 U/ml and 320 U/ml as a CoPs against any severity of rotavirus disease, along with binomial 95% confidence intervals using the Wilson Interval.[14] Multivariable logistic regression was used to assess the relationship between vaccine eligibility, age and attainment of the two thresholds for CoP in separate models for children under the age of 7 years (maximum age for vaccine-eligible cohort).

In the vaccine-ineligible population, some of the samples were collected in the vaccine era. We therefore conducted sensitivity analyses for all methods, which removed samples that came from the vaccine-ineligible population that were collected in the vaccine era. All analyses were conducted in RStudio version 2024.09.1+394 with R, version R-4.4.1.[23]

## Results

At total of 393 serum samples were from vaccine-eligible participants, and the remaining 807 were from vaccine-ineligible participants (Table 1.). In the vaccine-ineligible population, 288 samples were collected in the vaccine era. A total of 1200 samples were tested for anti-rotavirus IgG. Fewer samples (1108) could be tested for anti-rotavirus IgA as some samples had been exhausted. A total of 1009 IgA and 1054 IgG test results were valid, after further dilutions.

**Table 1.** Participant characteristics by vaccine eligibility.

|  | Vaccine-ineligible |  |  | Vaccine-eligible<br>(n=393) |
| --- | --- | --- | --- | --- |
|  | Sample taken pre-vaccine era (n=519) | Sample collected vaccine era (n=288) | Total vaccine-ineligible (n=807) |  |
| **Sex (%)** |  |  |  |  |
| Female | 222 (42.8) | 139 (48.3) | 361 (44.7) | 201 (51.1) |
| Male | 291 (56.1) | 149 (51.7) | 440 (54.5) | 191 (48.6) |
| Unknown | 6 (1.2) | 0 (0.0) | 6 (0.7) | 1 (0.3) |
| **Age group (%)** |  |  |  |  |
| <3m | 51 (9.8) | 0 (0.0) | 51 (6.3) | 55 (14.0) |
| 3-11m | 50 (9.6) | 4 (1.4) | 54 (6.7) | 42 (10.7) |
| 12-23m | 55 (10.6) | 22 (7.6) | 77 (9.5) | 71 (18.1) |
| 2y | 58 (11.2) | 29 (10.1) | 87 (10.8) | 63 (16.0) |
| 3y | 58 (11.2) | 31 (10.8) | 89 (11.0) | 61 (15.5) |
| 4y | 59 (11.4) | 26 (9.0) | 85 (10.5) | 65 (16.5) |
| 5-6y | 61 (11.8) | 53 (18.4) | 114 (14.1) | 36 (9.2) |
| 7-9y | 16 (3.1) | 17 (5.9) | 33 (4.1) | 0 (0.0) |
| 10-14y | 23 (4.4) | 20 (6.9) | 43 (5.3) | 0 (0.0) |
| 15-24y | 19 (3.7) | 19 (6.6) | 38 (4.7) | 0 (0.0) |
| 25-44y | 30 (5.8) | 30 (10.4) | 60 (7.4) | 0 (0.0) |
| 45-59y | 20 (3.9) | 18 (6.2) | 38 (4.7) | 0 (0.0) |
| 60+ | 19 (3.7) | 19 (6.6) | 38 (4.7) | 0 (0.0) |

### Anti-rotavirus IgA in serum

In the vaccine-ineligible population, anti-rotavirus IgA serum levels increase steeply (slope, 1.33 log(U/ml)/year; 95% CI, 0.67–2.63) from birth until 24.96 months of age (95% CI 16.08–42.82) (Figure 1; Table 2). After 24.96 months the slope is shallower (0.05 log(U/ml)/year; 95% CI, 0.04–0.05); but is still positive (Supplementary Figure S1). In the vaccine-eligible population, a steep increase was observed from 2 months of age (slope, 4.61 log(U/ml)/year; 95% CI, 2.54–14.52) until a single knot located at 9.24 months of age (95% CI 3.14–13.27), reaching a concentration of 40.12 U/ml (95% CI, 16.06-56.60). After 9.24 months of age, IgA levels plateaued (slope, 0.01 log(U/ml)/year; 95% CI, −0.12– −0.24), reaching a concentration of 43.11 U/ml (95% CI, 22.77–85.03) at age 7 years.

**Table 2.** Knot and slope parameter estimates from linear spline censored regression for anti-rotavirus IgA and IgG in vaccine-eligible and ineligible populations.

| Vaccine eligibility | Knots, age in months (95% CI) |  |  | Model slopes<sup>a</sup> in log(U/ml)/year (95% CI) |  |  |  |
| --- | --- | --- | --- | --- | --- | --- | --- |
|  | Knot 1 | Knot 2 | Knot 3 | Slope 1 | Slope 2 | Slope 3 | Slope 4 |
| **IgA** |  |  |  |  |  |  |  |
| Ineligible | 24.96 (16.08 to 42.82) | - | - | 1.33 (0.67 to 2.63) | 0.05 (0.04 to 0.05) | - | - |
| Eligible | 9.24 (3.14 to 13.27) | - | - | 4.61 (2.54 to 14.52) | 0.01 (-0.12 to 0.24) | - | - |
| **IgG** |  |  |  |  |  |  |  |
| Ineligible | 3.00 (2.82 to 3.38) | 14.56 (10.52 to 24.00) | 342.33 (92.44 to 417.61) | -15.48 (-18.96 to -13.32) | 2.34 (1.28 to 4.55) | 0.07 (0.05 to 0.25) | 0.00 (-0.01 to 0.02) |
| Eligible | 3.6 (1.52 to 9.96) | - | - | -6.17 (-13.2 to -1.84) | 0.17 (0.01 to 0.36) | - | - |

red text = positive slope; blue text = negative slope; black text = null slope
<sup>a</sup> slopes (m) are from a log-linear model $y = c^{e(mx)}$ , m therefore takes the units which are the inverse of the units of the x-axis, c = intercept, x = value of x (age in months), y = value of y (concentration in U/ml)

**Figure 1.**
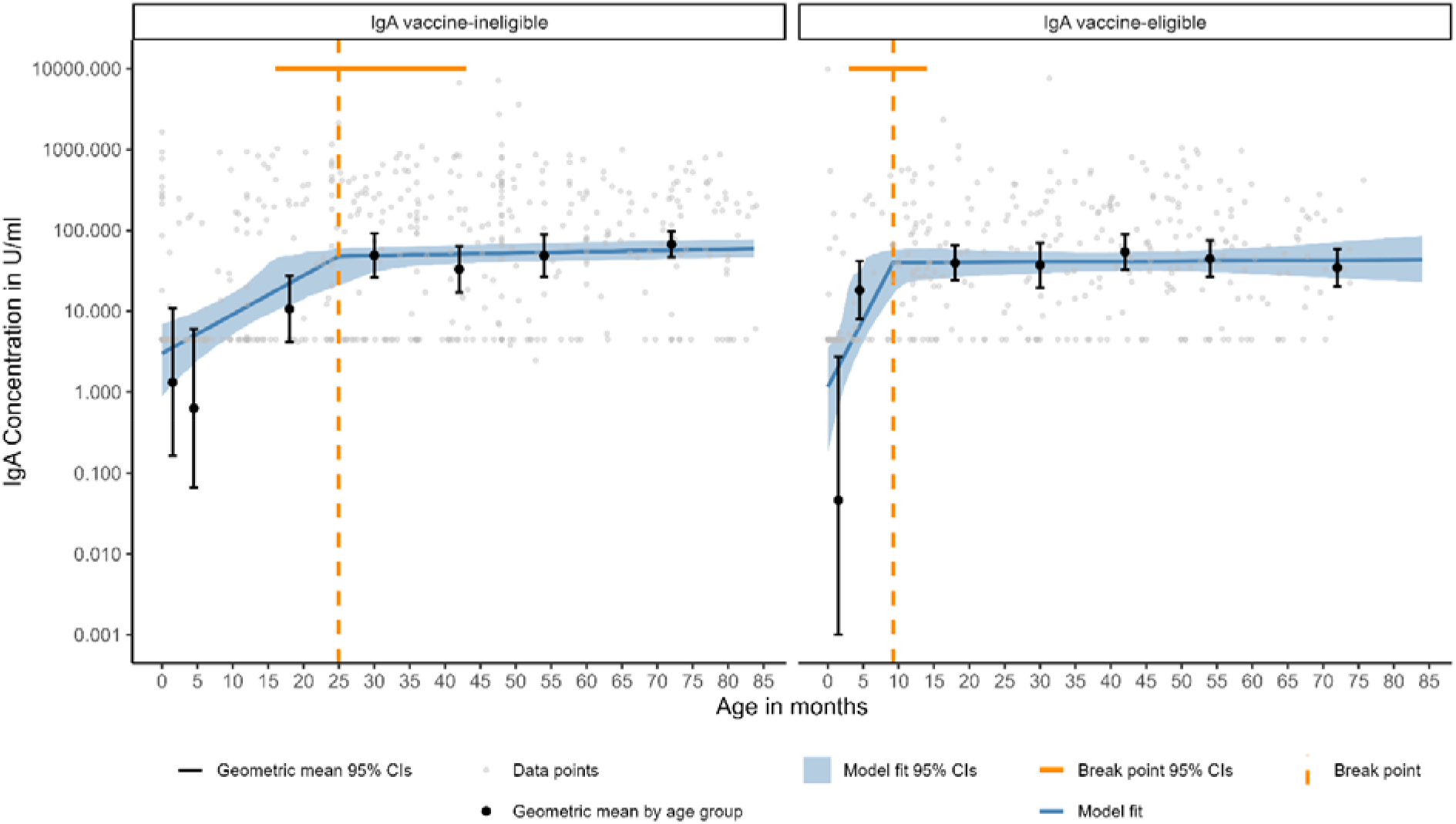
Linear spline censored regression model assessing the associations between age and anti-rotavirus IgA up to the age of 7 years in vaccine-eligible and vaccine-ineligible populations. Data below the lower limit of quantification (LLoQ) are plotted at the LLoQ.

As serum anti-rotavirus IgA has been used as an indicator of a CoP against disease, we assessed the proportion of IgA samples above a threshold of ≥20 U/ml,≥160 U/ml and ≥320 U/ml as age increased (Figure 2, supplementary Table S2). In the vaccine-eligible population, the proportion of samples ≥20 U/ml (CoP against severe disease) was higher in the 12-23 month age group but otherwise comparable to the vaccine-ineligible population at other ages. The proportion of samples with serum anti-rotavirus IgA ≥160 U/ml (CoP against rotavirus disease of any severity) was lower in the vaccine-eligible population. In the vaccine-ineligible population, the proportion reaching this threshold increased dramatically in age groups 15 years and older to 0.81 (0.65 to 0.91) in 60+ year-olds. In logistic regression analyses adjusted for age (supplementary Table S3), the vaccine-eligible population had lower odds of attaining a threshold of IgA ≥160 U/ml and when compared to the vaccine-ineligible population (adjusted odds ratio [aOR], 0.66; 95% CI, 0.48–0.91), for a threshold of IgA ≥20 U/ml the effect was in the opposite direction, although this was not significant (aOR, 1.29; 95% CI, 0.95–1.75). The proportion of samples with serum anti-rotavirus IgA ≥320 U/ml are lower but show a similar pattern to ≥160 U/ml.

**Figure 2.**
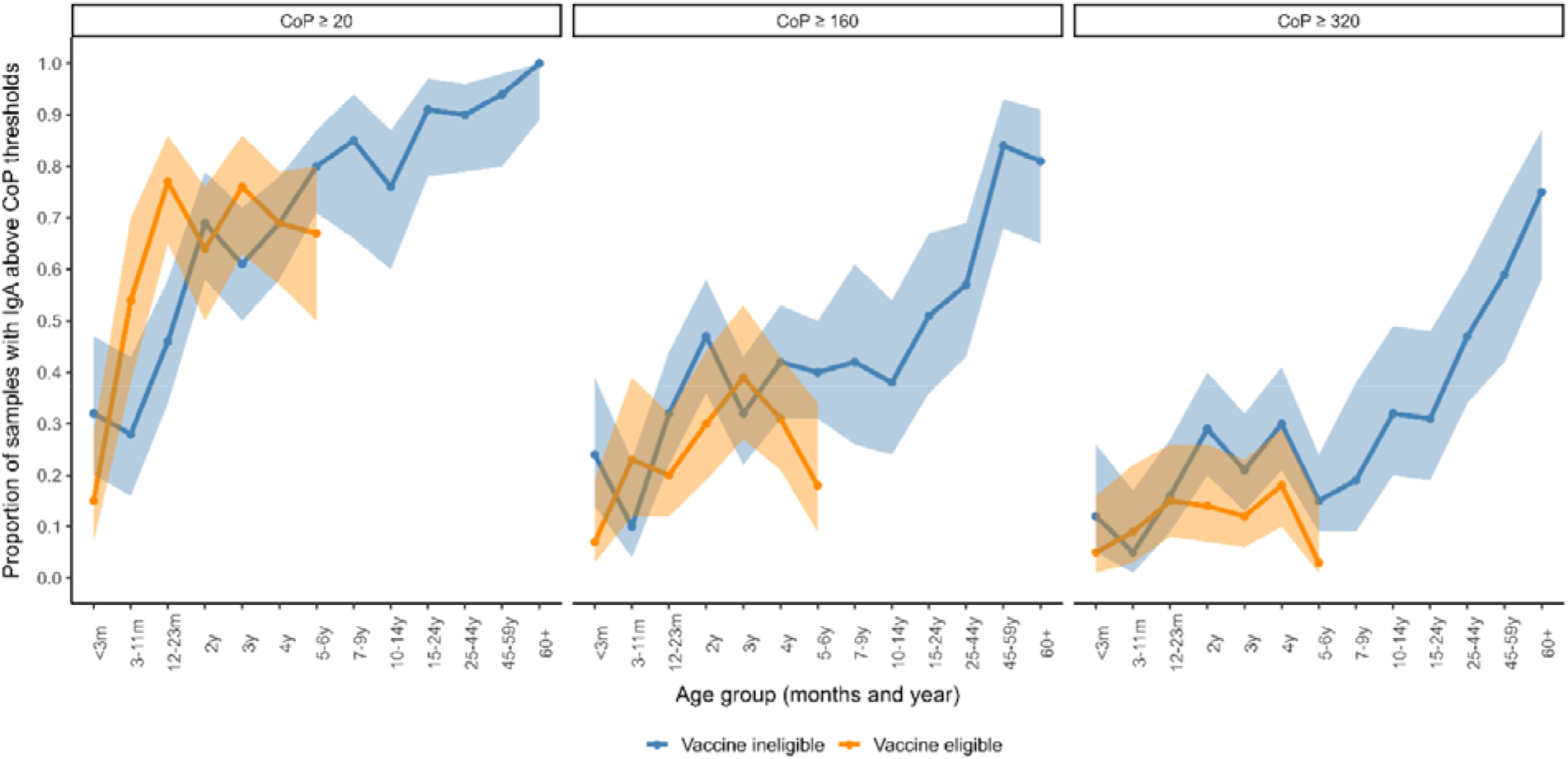
The proportion of samples that attained anti-rotavirus IgA CoP thresholds indicative of being protected against severe rotavirus disease (≥20 U/ml) or rotavirus disease of any severity (≥160 U/ml) by age group and vaccine eligibility of population. Age-specific point estimates and confidence intervals are included in supplementary Table S2.

**Figure 3.**
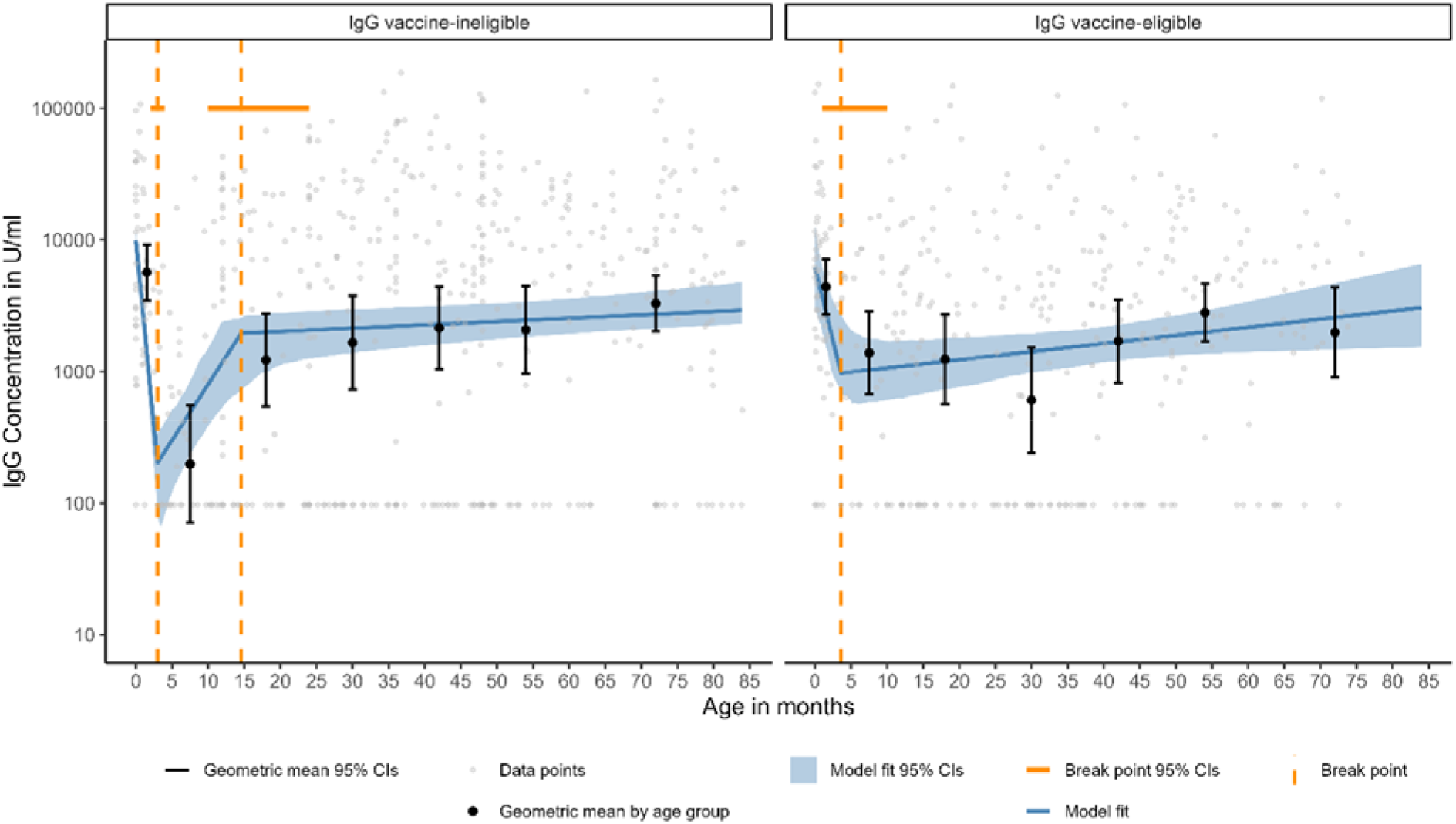
Linear spline censored regression model assessing the associations between age and anti-rotavirus IgG up to the age of 7 years in vaccine-eligible and vaccine-ineligible populations

### Anti-rotavirus IgG in serum

For anti-rotavirus IgG, maternal antibodies are likely detected in the vaccine-ineligible and eligible populations before 3 months of age (Figure 1). In the vaccine-ineligible population, IgG waning occurs after 2 months of age and reaches a low of 204.07 U/ml (95% CI, 88.32–342.62) at age 3.0 (95% CI, 2.8-3.4) months. In the vaccine-eligible population, waning is detected, to a low of 974.03 U/ml (95% CI, 676.25-2731.10) at 3.6 months of age, but not to the level seen in the vaccine-ineligible population likely due to immune boosting via vaccination with one or two doses between 2-5 months of age. In the vaccine-eligible cohort between 3-11 months of age, IgG levels are 7 times greater (GMC, 1388.67 U/mL; 95% CI, 675.03-2856.78) than those induced by natural exposure in the vaccine-ineligible cohort (GMC, 198.54 U/ml; 95% CI, 71.31-552.78) (Supplementary Table S1).

In the vaccine-ineligible cohort, IgG increases from age 3 months to age 14.6 months (slope, 2.34 log(U/ml)/year; 95% CI, 1.28-4.55) (Table 2 and Supplementary Figure S2), then there is a shallower increase in IgG to age 28.5 years. This pattern is not seen in the vaccine-eligible population, where IgG levels slightly increase between 3.6 months and 7 years of age, at a rate of 0.17 log(U/ml)/year (95% CI, 0.01-0.36).

### Sensitivity analyses

Restricting analyses in the vaccine-ineligible cohort to only those with a sample collected in the pre-vaccine era had minor effects on estimates for CoP analyses (supplementary Table S3). The only variation in the findings was that the knot for IgG at 28.5 years was dropped, but like the main findings, IgA and IgG levels continued to increase through childhood into adulthood (supplementary Table S4). All other sensitivity analysis findings remained consistent with the main analyses.

## Discussion

For the first time in any HIC, this seroepidemiological study has shown that in the pre-vaccine era natural immunity to rotavirus increases with age, presumably due to boosting events in adulthood and later life. In the vaccine-eligible cohort, we showed that levels of anti-rotavirus IgA and IgG antibodies are greater than that induced by natural exposure between 2 months and 24 months of age, and we inferred that protection against severe disease is comparable in the vaccine-eligible and vaccine-ineligible populations between 2 and 7 years of age. However, anti-rotavirus IgA antibody levels plateaued and IgG levels only mildly increased after infancy in the vaccine-eligible population, and inferred protection against rotavirus disease of any severity is lower among vaccine-eligible children.

Our findings in the vaccine-ineligible population are similar to a pre-vaccine cohort study in Brazil, where IgG and IgA seropositivity accumulates with age as a result of natural exposure and re-infections.[11] Birth cohort studies in India and Mexico have shown that natural rotavirus infections were associated with protection against disease but provided lesser protection against reinfection.[24,25] Reinfections accumulate as children age into adulthood and the majority of individuals experience multiple infections, with subsequent infections milder or asymptomatic after two consecutive infections.[24] In contrast, we did not observe continued accumulation of rotavirus-specific IgA and IgG in our vaccine-eligible population after the age of vaccination, and we found a lower proportion of individuals with anti-rotavirus IgA >=160 U/ml and IgA >=320 U/ml, indicative of lesser protection against milder disease, compared to the vaccine-ineligible cohort.

Our finding in the vaccine-eligible population follow the rapid and dramatic reduction in rotavirus disease after vaccine introduction in the UK in 2013.[5,6] This likely indicates that the force of infection and exposure to rotavirus infections in the vaccine era are less frequent, in keeping with reports of indirect vaccine effects in HICs.[5,7,26,27] Potential explanations for these findings, which are not necessarily mutually exclusive, include: 1. due to vaccine-induced protection, natural infections are not eliciting substantial immune boosting events; 2. the frequency of natural infection is too modest to boost immunity beyond that of vaccine-induced immunity; and, 3. waning of vaccine-induced immunity is being countered by natural infections/re-infections, leaving a plateau in rotavirus-specific IgA and a milder increase in IgG levels between 12 months and 6 years of age.

Our findings have implications for public health, and health protection research. Reduced or waning immunity due to less natural exposure to rotavirus infection could render older children and adults vulnerable to sporadic outbreaks and increased incidence of rotavirus disease. Since there is an age- associated relationship with severity of diarrhoeal disease, milder disease in older children and healthy adults is more likely to be self-limiting with presentation at primary care, emergency care or digital triage, rather than inpatient hospital settings. For example, a US study showed increases in emergency department visits for diarrhoea in older children 5-12 years of age since rotavirus vaccine introduction.[28] However, it should be noted that if there is substantial waning of antibodies, then vulnerable older adults may be more susceptible to severe disease outcome. Since testing for rotavirus in the UK is primarily recommended for children under five years attending healthcare with severe gastroenteritis,[29] changes in clinical presentation may have implications for diagnostic testing algorithms. Therefore, the societal, economic and healthcare use associated with an altered epidemiological and clinical severity profile of rotavirus disease will need to be assessed, using robust applied epidemiology of healthcare and community disease data. Finally. results from our study will be used to update rotavirus transmission model parameters on the susceptibility to infection and protection against disease in HICs.[30–32]

### Limitations

The data did not include information on individual rotavirus vaccination status. However, evidence shows that the majority (~94%) of the vaccine-eligible cohort would be vaccinated.[33–35] The data are also, cross-sectional, so age-related changes in immunity are at the population level not the individual level. Data are also subject to temporal confounding unrelated to vaccination and do not contain any clinical or microbiological evidence of rotavirus infection or disease. Our model assumptions of linear trends by age were selected for mathematical convenience rather than biological plausibility. Further research is required to investigate whether immunity and protection in the vaccine-eligible population remains static or wanes as vaccinated cohorts reach secondary school age. Lastly, the samples included in the study are from a convenience sample and though representative of the healthy general population, the reason for diagnostic testing of the original sample is unknown.[17]

## Conclusions

For first time in a vaccine-naïve UK population, we show antibody-based immunity to rotavirus at the population level increases through childhood and into adulthood. However, in the vaccine era, in the context of reduced rotavirus disease and transmission, rotavirus immunity does not continue to increase with age after vaccine-induced immune exposure. There is also evidence through CoP threshold analysis that in the vaccine era there is likely lower protection against milder disease. This indicates that there is reduced infection exposure and immune boosting opportunities, which could result in shifts in rotavirus disease severity and age of onset, to milder disease in older paediatric populations. Further research is needed to understand disease dynamics, rotavirus disease profiles and economic costs as vaccinated cohorts age.

## Data Availability

Deidentified aggregated data used for analysis can be made available on request, with valid study protocol and approvals. Additional study documents including the study protocol are available on request to the corresponding author.

## Acknowledgements

We thank Professor Ray Borrow, Dr Simon Tonge and Dr Ezra Linley from the Vaccine Evaluation Unit for assisting with study approval and selecting the serum samples from the UKHSA Seroepidemiology Unit. Dr Merryn Voysey from University of Oxford for statistical advice related to the study protocol.

## Contributions

DH, VEP and MIG conceived the study. DH, MIG, and VEP designed the study, with contributions from NF, CB and NC. KCJ, MIG, JD, JM, KR oversaw laboratory activities. DH, JD, KCJ, MIG, and KR oversaw data curation. MH and VEP provided statistical support. DH completed formal analysis. JD, DH, KR and KCJ have directly accessed and verified the underlying data reported in the manuscript. DH and MIG jointly wrote the first draft of the manuscript. All authors have contributed to data interpretation while editing and commenting on the draft manuscript. All authors have read and approved the final manuscript. DH, had final responsibility for the decision to submit for publication.

