## supplementary for "Age-specific immunity to rotavirus infection and the risk of disease before and after rotavirus vaccine introduction in the United Kingdom: an observational, seroepidemiological study"

### Additional figures and tables supplementing main findings

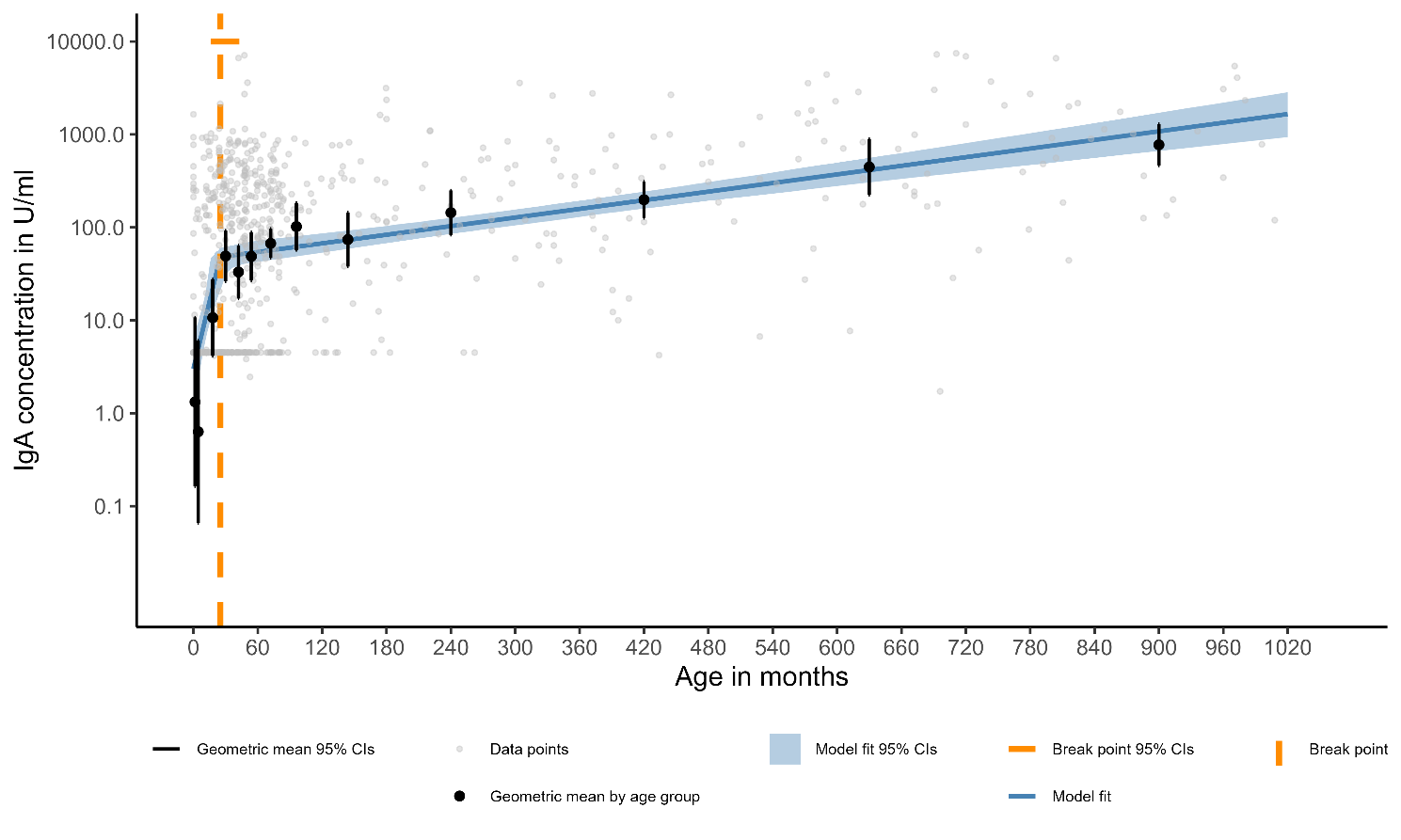

**Figure S1. Linear spline censored regression model assessing the associations between age and anti-rotavirus IgA for all ages in the vaccine-ineligible population**

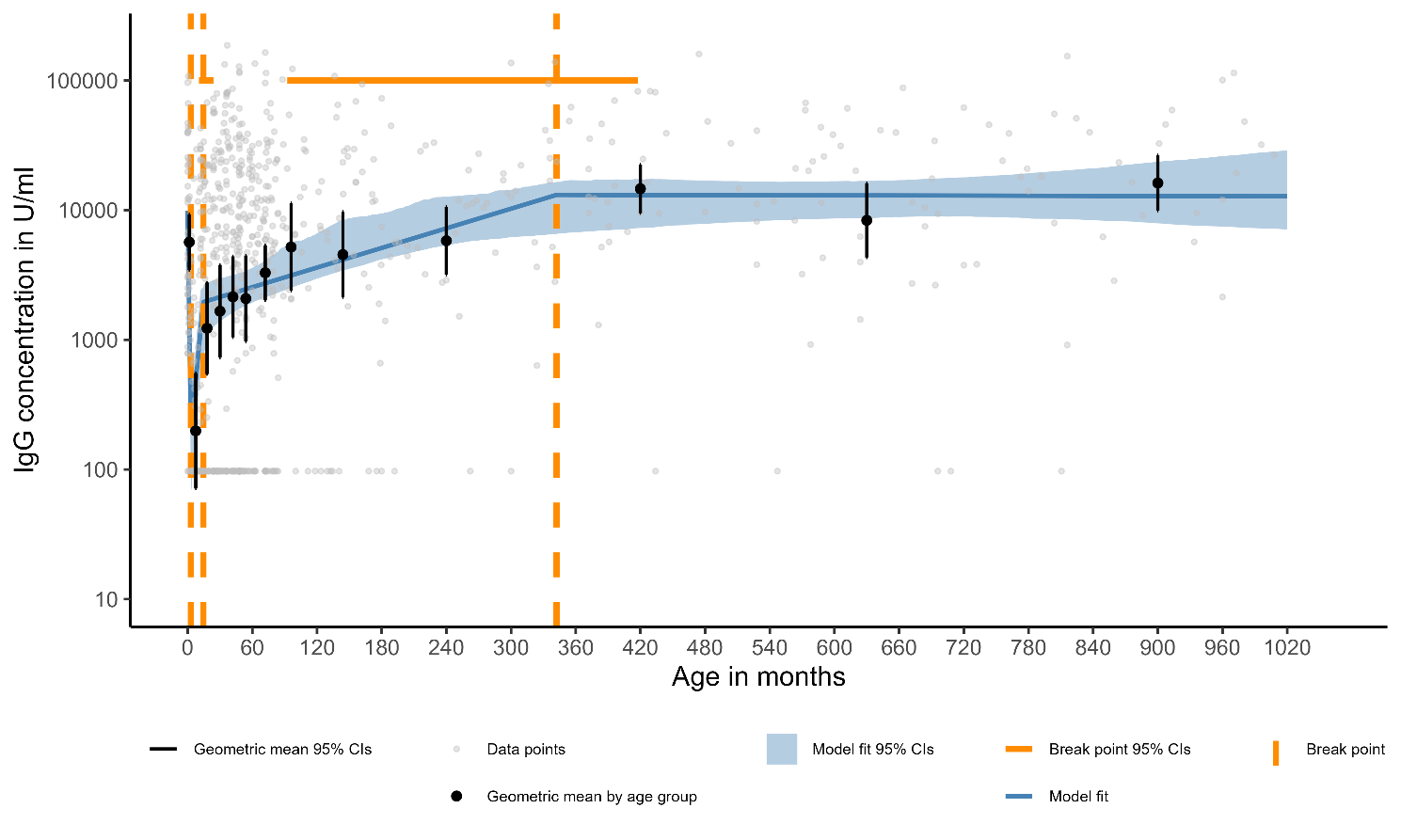

**Figure S2. Linear spline censored regression model assessing the associations between age and anti-rotavirus IgG for all ages in the vaccine-ineligible population**

**Table S1. Geometric mean concentrations of anti-rotavirus IgA and IgG by age group and vaccine eligibility of population**

| **Age group** | **IgA geometric mean concentration in U/ml (95% CI)** | | **IgG geometric mean concentration in U/ml (95% CI)** | |
| --- | --- | --- | --- | --- |
|  | **Vaccine-ineligible** | **Vaccine-eligible** | **Vaccine-ineligible** | **Vaccine-eligible** |
| <3m | 1.33 (0.16 to 10.74) | 0.05 (0 to 2.76) | 5665.54 (3461.42 to 9273.18) | 4411.6 (2720.88 to 7152.91) |
| 3-11m | 0.63 (0.07 to 6.05) | 18.23 (8.01 to 41.53) | 198.54 (71.31 to 552.78) | 1388.67 (675.03 to 2856.78) |
| 12-23m | 10.66 (4.14 to 27.43) | 39.58 (24.13 to 64.92) | 1226.99 (544.85 to 2763.18) | 1241.95 (568.4 to 2713.67) |
| 2y | 49.09 (26.16 to 92.1) | 37.07 (19.6 to 70.09) | 1662 (730.76 to 3779.97) | 608.23 (243.02 to 1522.31) |
| 3y | 33.08 (17.11 to 63.97) | 54.13 (32.7 to 89.62) | 2145.5 (1044.29 to 4407.94) | 1710.13 (827.12 to 3535.82) |
| 4y | 48.67 (26.76 to 88.52) | 44.61 (26.58 to 74.88) | 2080.1 (968.12 to 4469.29) | 2811.66 (1699.6 to 4651.33) |
| 5-6y | 67.18 (46.8 to 96.43) | 34.6 (20.35 to 58.83) | 3288.89 (2017.01 to 5362.79) | 1988.86 (904.58 to 4372.84) |
| 7-9y | 101.87 (57.13 to 181.65) | NA | 5195.95 (2385.48 to 11317.6) | NA |
| 10-14y | 73.88 (37.85 to 144.2) | NA | 4541.43 (2119.55 to 9730.64) | NA |
| 15-24y | 143.87 (83.02 to 249.32) | NA | 5804.99 (3185.22 to 10579.46) | NA |
| 25-44y | 198.07 (126.14 to 311.02) | NA | 14624.03 (9475.36 to 22570.35) | NA |
| 45-59y | 446.56 (223.33 to 892.91) | NA | 8344.49 (4314.49 to 16138.76) | NA |
| 60+y | 772.96 (463.2 to 1289.85) | NA | 16189.03 (9859.35 to 26582.36) | NA |

**Table S2. Proportion of IgA samples attaining correlates of protection thresholds indicative of protection against severe rotavirus disease (≥20 U/ml) and rotavirus disease of any severity (≥160 U/ml) by age group and vaccine eligibility of population**

| **Age group** | **Proportion of IgA samples ≥20 U/ml (95% CI)** | | **Proportion of IgA samples ≥160 U/ml (95% CI)** | | **Proportion of IgA samples ≥320 U/ml (95% CI)** | |
| --- | --- | --- | --- | --- | --- | --- |
|  | **Vaccine-ineligible** | **Vaccine-eligible** | **Vaccine-ineligible** | **Vaccine-eligible** | **Vaccine-ineligible** | **Vaccine-eligible** |
| <3m | 0.32 (0.2 to 0.47) | 0.15 (0.07 to 0.28) | 0.24 (0.14 to 0.39) | 0.07 (0.03 to 0.19) | 0.12 (0.05 to 0.26) | 0.05 (0.01 to 0.16) |
| 3-11m | 0.28 (0.16 to 0.43) | 0.54 (0.38 to 0.7) | 0.1 (0.04 to 0.23) | 0.23 (0.12 to 0.39) | 0.05 (0.01 to 0.17) | 0.09 (0.03 to 0.22) |
| 12-23m | 0.46 (0.34 to 0.58) | 0.77 (0.65 to 0.86) | 0.32 (0.22 to 0.44) | 0.2 (0.12 to 0.32) | 0.16 (0.09 to 0.27) | 0.15 (0.08 to 0.26) |
| 2y | 0.69 (0.58 to 0.79) | 0.64 (0.5 to 0.76) | 0.47 (0.36 to 0.58) | 0.3 (0.19 to 0.44) | 0.29 (0.2 to 0.4) | 0.14 (0.07 to 0.26) |
| 3y | 0.61 (0.5 to 0.72) | 0.76 (0.63 to 0.86) | 0.32 (0.22 to 0.43) | 0.39 (0.27 to 0.53) | 0.21 (0.13 to 0.32) | 0.12 (0.06 to 0.23) |
| 4y | 0.69 (0.58 to 0.78) | 0.69 (0.57 to 0.79) | 0.42 (0.31 to 0.53) | 0.31 (0.21 to 0.43) | 0.3 (0.21 to 0.41) | 0.18 (0.1 to 0.29) |
| 5-6y | 0.8 (0.71 to 0.87) | 0.67 (0.5 to 0.8) | 0.4 (0.31 to 0.5) | 0.18 (0.09 to 0.34) | 0.15 (0.09 to 0.24) | 0.03 (0.01 to 0.15) |
| 7-9y | 0.85 (0.66 to 0.94) | NA | 0.42 (0.26 to 0.61) | NA | 0.19 (0.09 to 0.38) | NA |
| 10-14y | 0.76 (0.6 to 0.87) | NA | 0.38 (0.24 to 0.54) | NA | 0.32 (0.2 to 0.49) | NA |
| 15-24y | 0.91 (0.78 to 0.97) | NA | 0.51 (0.36 to 0.67) | NA | 0.31 (0.19 to 0.48) | NA |
| 25-44y | 0.9 (0.79 to 0.96) | NA | 0.57 (0.43 to 0.69) | NA | 0.47 (0.34 to 0.6) | NA |
| 45-59y | 0.94 (0.8 to 0.98) | NA | 0.84 (0.68 to 0.93) | NA | 0.59 (0.42 to 0.74) | NA |
| 60+ | 1 (0.89 to 1) | NA | 0.81 (0.65 to 0.91) | NA | 0.75 (0.58 to 0.87) | NA |

### Sensitivity analyses described in the main paper

**Table S3. Association between vaccine eligibility and the proportion of IgA samples attaining correlates of protection thresholds indicative of protection against severe rotavirus disease (≥20 U/ml) and rotavirus disease of any severity (≥160 U/ml and (≥320 U/ml) in children under the age of 7 years**

| **Analysis** | **CoP IgA ≥20 U/ml** | | | **CoP IgA ≥160 U/ml** | | | **CoP IgA ≥320 U/ml** | | |
| --- | --- | --- | --- | --- | --- | --- | --- | --- | --- |
|  | **Vaccine-ineligible**  **(%)** | **Vaccine-eligible**  **(%)** | **aOR**  **(95% CI)** | **Vaccine-ineligible**  **(%)** | **Vaccine-eligible**  **(%)** | **aOR**  **(95% CI)** | **Vaccine-ineligible**  **(%)** | **Vaccine-eligible**  **(%)** | **aOR**  **(95% CI)** |
| Original | 279/464 (60.1) | 207/332 (62.3) | 1.29 (0.95 to 1.75) | 163/464 (35.1) | 83/332 (0.25) | 0.66 (0.48 to 0.91) | 91/464 (19.6) | 39/332 (11.7) | 0.56 (0.37 to 0.84) |
| Sensitivity* | 188/322 (58.4) | 207/332 (62.3) | 1.23 (0.89 to 1.71) | 108/322 (33.5) | 83/332 (0.25) | 0.67 (0.48 to 0.94) | 59/322 (18.3) | 39/332 (11.7) | 0.60 (0.38 to 0.92) |

*(vaccine-ineligible population only includes those with a sample collected in the pre-vaccine era)

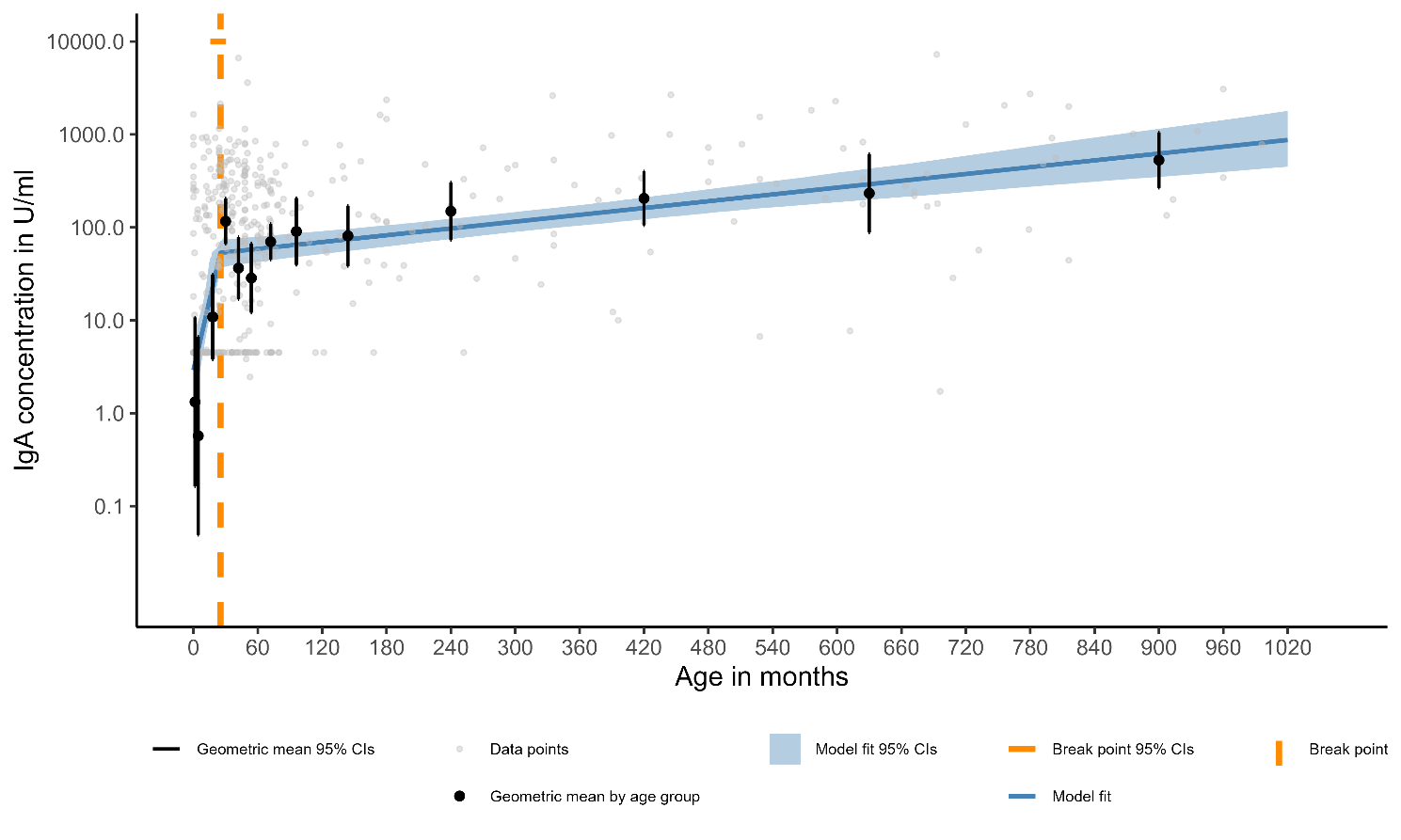

**Figure S3. Linear spline censored regression model assessing the associations between age and anti-rotavirus IgA for all ages in the vaccine-ineligible, with sample collected in the pre-vaccine era only**

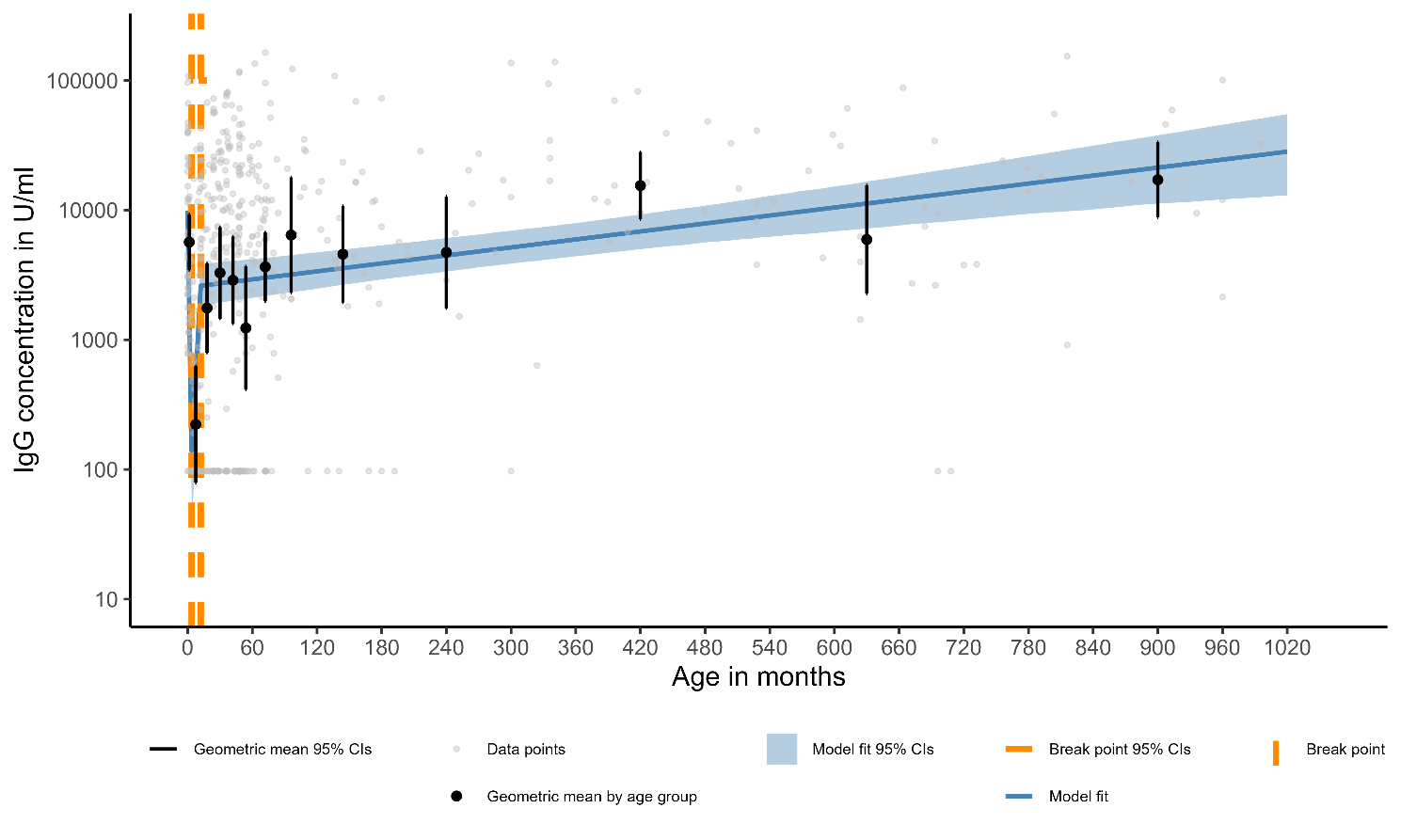
**Figure S4. Linear spline censored regression model assessing the associations between age and anti-rotavirus IgG for all ages in the vaccine-ineligible, with sample collected in the pre-vaccine era only**

**Table S4. Knot and slope parameter estimates from linear spline censored regression for anti-rotavirus IgA and IgG in all vaccine-ineligible compared to models which only include samples from vaccine-ineligible collected in pre-vaccine era**

| Vaccine ineligibility | Knots in age in months (95% CI) | | | Model slopes ^ⴕ^ in log(U/ml)/year (95% CI) | | | |
| --- | --- | --- | --- | --- | --- | --- | --- |
|  | Knot 1 | Knot 2 | Knot 3 | Slope 1 | Slope 2 | Slope 3 | Slope 4 |
| IgA | | | | | | | |
| All samples | 24.96 (16.08 to 42.82) |  | - | 1.33 (0.67 to 2.63) | 0.05 (0.04 to 0.05) |  |  |
| Sample collected in pre-vaccine era only | 25.20 (15.71 to 30.48) |  |  | 1.38 (0.90 to 2.53) | 0.04 (0.02 to 0.05) |  |  |
| IgG | | | | | | | |
| All samples | 3.00 (2.82 to 3.38) | 14.56 (10.52 to 24.00) | 342.33 (92.44 to 417.61) | -15.48 (-18.96 to -13.32) | 2.34 (1.28 to 4.55) | 0.07 (0.05 to 0.25) | 0.00 (-0.01 to 0.02) |
| Sample collected in pre-vaccine era only | 3.60 (2.74 to 5.30) | 12.17 (10.07 to 18.00) |  | -14.28 (-17.76 to -9.01) | 4.13 (1.96 to 7.37) | 0.02 (0.01 to 0.04) |  |

red text = positive slope; blue text = negative slope; black text = null slope

^ⴕ^ slopes (m) are from a log-linear model $y=c^{e(\mathrm{mx})}$, m therefore takes the units which are the inverse of the units of x-axis, c = intercept, x = value of x (age in months), y = value of y (concentration in U/ml)
